# Applying models of care for total hip and knee arthroplasty: External validation of a published predictive model to identify extended stay risk prior to lower-limb arthroplasty

**DOI:** 10.1101/2023.01.12.23284462

**Authors:** Meredith Harrison-Brown, Corey Scholes, Milad Ebrahimi, Christopher Bell, Garry Kirwan

**Affiliations:** Department of Orthopaedics, QEII Jubilee Hospital, Brisbane, QLD, Australia; EBM Analytics, Sydney, NSW, Australia; Faculty of Medicine, The University of Queensland, Brisbane, Qld, Australia; Department of Physiotherapy, QEII Jubilee Hospital, Brisbane, QLD, Australia; School of Allied Health Sciences, Griffith University, Gold Coast, Qld, Australia

**Keywords:** Joint arthroplasty, model of care, extended length of stay, prediction algorithm

## Abstract

**Introduction/Aims:** Multiple predictive tools have been developed to identify patients requiring an extended hospital stay following lower limb arthroplasty. Use at new sites requires verification of appropriate data coverage and evidence of validity in a new population. The aim of this study was to externally validate a previously reported model for identifying patients requiring an extended (5+ day) stay following total hip or knee replacement in a medium-sized public hospital orthopaedic department.

**Methods:** Electronic medical records were accessed and retrospective data extracted from 200 randomly selected total hip or knee arthroplasty patients. Data fields were matched to the candidate model and organised for validation analysis. Model validation was assessed with model discrimination, calibration on both original (unadjusted) and adjusted forms of the candidate model. Decision curve analysis was conducted on the outputs of the adjusted model.

**Results:** The original model performed poorly in the validation dataset, grossly overestimating length of stay. Performance improved following adjustment of the model intercept and model coefficients, although the model remained poorly calibrated at low and medium risk threshold and net benefit of the adjusted model was modest.

**Conclusion:** Consideration of predictive models to improve resource allocation for postoperative management of arthroplasty patients should include practical assessment of discrimination, calibration and net benefit of intervention at a clinically acceptable threshold prior to implementation within their local setting. This forms a clinically meaningful assessment of the accuracy, as well as costs and benefits associated with the desired change in practice. The selection of an appropriate model for quality care improvement remains challenging.

## Introduction

The volume of lower limb (hip and knee) total joint arthroplasty (TJA) was steadily increasing prior to the Covid19 pandemic. Following suspension of elective surgery in some Australian jurisdictions as a result of the pandemic, the expectation is that this demand will substantially grow in the coming years. The Australian Orthopaedic Association National Joint Replacement Registry Annual Report (2021) [1] states there were 92,619 primary total hip and knee replacements performed in 2020. Interestingly, despite prolonged lockdowns and pauses on elective surgeries, surgical volume was only minimally affected, at approximately 3.9% lower than 2019 volumes [2]. However, the prolonged effects for future surgical waitlists from 2022 and beyond are unknown. Based on current Australian population growth and TJA growth rates, the number of joint replacements is projected to undergo unsustainable growth of 276% and 208% for knee and hip replacements, respectively over the next 10 years [3]. As a result, the healthcare system is under significant pressure to reduce the costs associated with these procedures in order to meet the demand. Current evidence suggests that length of hospital stay is the primary measure that correlates to financial burden [4–6], therefore this metric has become a key target for improvement.

Although most patients undergoing lower limb arthroplasty can safely follow a relatively short stay postoperative protocol, certain patients are at risk of complications or suboptimal clinical improvement necessitating a prolonged length of stay (LOS) [5,7–11]. A key factor to helping reduce the risk of prolonged LOS is the ability to accurately identify at-risk patients prior to surgery [5,7,11]. With accurate identification, a multi-factor approach can be adopted to tailor interventions to individual needs at both the pre-operative and post-operative stage. By doing so, modifiable risk factors can be addressed and patient outcomes optimised from the acute through to the subacute phase of recovery [8,11–13].

Numerous models to predict LOS in lower limb joint arthroplasty have been published [14]. Large clinical centres often develop and internally validate their own predictive models for use in their institution. However, very few are externally validated and many are reported too poorly to be replicated independently [15,16]. Furthermore, facilities can be constrained by the time required to generate sufficient sample size, introducing the risk of process changes over time (e.g. perioperative management, patient selection) that impact on the underlying data structure. The broad introduction of enhanced recovery after surgery programs is a key example of how system wide change will inherently alter the accuracy of a predictive tool. Therefore, to efficiently prioritise patient care based on risk of extended hospital stay, it is necessary to implement models and tools developed elsewhere. However, this process can be fraught without careful consideration for the appropriateness of the selected tool within the clinical environment and patient population (case-mix) [17]. A process of validation external to the setting in which the original model/tool was developed is necessary to mitigate the risk of applying a tool that lacks discrimination or is poorly calibrated for the new patient population.

A number of tools have been reported in the literature to effectively predict patients at risk of extended LOS [11–13,18]. Oldmeadow et al (2003), first developed the Risk Assessment and Prediction Tool (RAPT) demonstrating a 75% accuracy in predicting the need for extended rehabilitation. Sconza et al (2019) confirmed a similar finding when conducting a systematic review on studies investigating the RAPT. Other predictive models such as those developed by Winemaker et al (2015) or Barsoum et al 2010, report similar levels of accuracy and thus implies that predictive tools are an effective strategy for clinicians to implement into clinical practice. However, which tool best fits an individual facility or a particular model of care? Each tool uses a wide range of differing variables, and individual health facilities collect a varied range of patient data based on local practices. In a typically pragmatic approach to health care, predictive tools are often implemented and inevitably modified to fit the needs of the service. However, there is limited evidence to show that such tools can be applied indiscriminately in this way across all clinical settings [8,11–13,18,19].

We have chosen a model [18] based on the best compatibility of the predictor set against variables collected routinely at our service. The present study aims to identify and externally validate a predictive model for classifying patients at risk of an extended hospital stay (5+ days) after hip or knee TJA in a medium-sized public hospital, to assess its transportability and clinical utility for initiating earlier postoperative planning.

## Methods

### Study design - Source of data

This study is an external validation of a predictive model with a randomly selected subset of medical records in a retrospective cohort design. The study was reported according to the RECORD guidelines for studies conducted using routinely collected health data [20] and the TRIPOD guidelines for reporting of prognostic models [16]. The present study is a Type 4 analysis as per published guidelines [16]. Data was collected as part of standard medical records compilation for lower limb TJA performed 2-Feb-16 to 4-Apr-19. Medical records were entered by hospital staff directly into the integrated electronic medical record (iEMR) (Cerner, USA) introduced to the public health system within the state as of Nov-2018 as part of a digital hospital rollout, with the specific hospital converted in Apr 2019. The iEMR records were accessed for data collection purposes between Sep-19 and Feb-20. Prior to data collection, a study core dataset was developed, combining all outcome and predictor variables from the selected model as well as key indicators for patient data linkage. The research group comprised a collection team, led by orthopaedic physiotherapists and a quality team, external to the hospital. The core dataset was agreed by both teams following a pilot study to assess data feasibility, and then converted to a master database for construction through data linkage, chart review, as well as variable calculations and recoding. Data linkage for eligible patients was performed between exports from different hospital systems provided by the hospital’s decision support unit using unique patient identifiers (unit record number; URN) for exact matches between datasets. Date and time of admission as well as discharge were extracted from the data linkage process, and used to calculate LOS. Two reviewers performed a manual chart review of the patient’s electronic medical record for the remaining data items.

#### Data quality

Data quality was assessed during the data collection period by export of the mastersheet weekly and imported into Matlab (v2018b, Mathworks Inc, USA) for further processing. A script was used to i) check for data completeness relative to the records marked *completed* by the collection team and ii) check that responses matched the core dataset framework, iii) check for outliers in the continuous variables and iv) assess consistency between variables for accurate patient inclusion/exclusions. Discrepancies were highlighted to the collection team through a dedicated column (*discrepancies*) in the mastersheet and confirmed with additional record review prior to model/tool adaptation and validation.

#### Bias

Selection and information bias associated with data availability relative to the length of time from changeover to iEMR was mitigated by using random selection from the pool of all eligible records, rather than reverse chronological order. Random permutation [21] between 1 and 200 was used to select the final list of 200 patients with a random number generator function, set to zero seeds (Matlab 2018b, Mathworks Inc, USA).

#### Setting

The study is a single-centre analysis with 14 consultant surgeons operating within an orthopaedic department of a medium-size metropolitan public hospital located in South-East Queensland, Australia. Of the 200 randomly selected cases, a majority of the procedures were performed by 4 surgeons with 38, 34, 29 and 25 procedures respectively accounting for greater than 60% of surgeries. For the remaining TJA procedures, 4 surgeons accounted for 50 (25%) procedures with an additional 6 surgeons performing 10 or fewer procedures each.

### Participants

The iEMR was queried for all elective total hip and knee arthroplasties (no exclusions for revisions or bilateral surgeries) performed within the department between the dates listed. The results of the iEMR query were de-identified by removing patient names, addresses, and contact information. The query returned 1215 cases forming the database population **(Figure 1**). The available pool of perioperative records was limited to those that were raised following the hospital transition to iEMR in April 2019, leaving a pool of 331 patient records.

**Figure 1.**
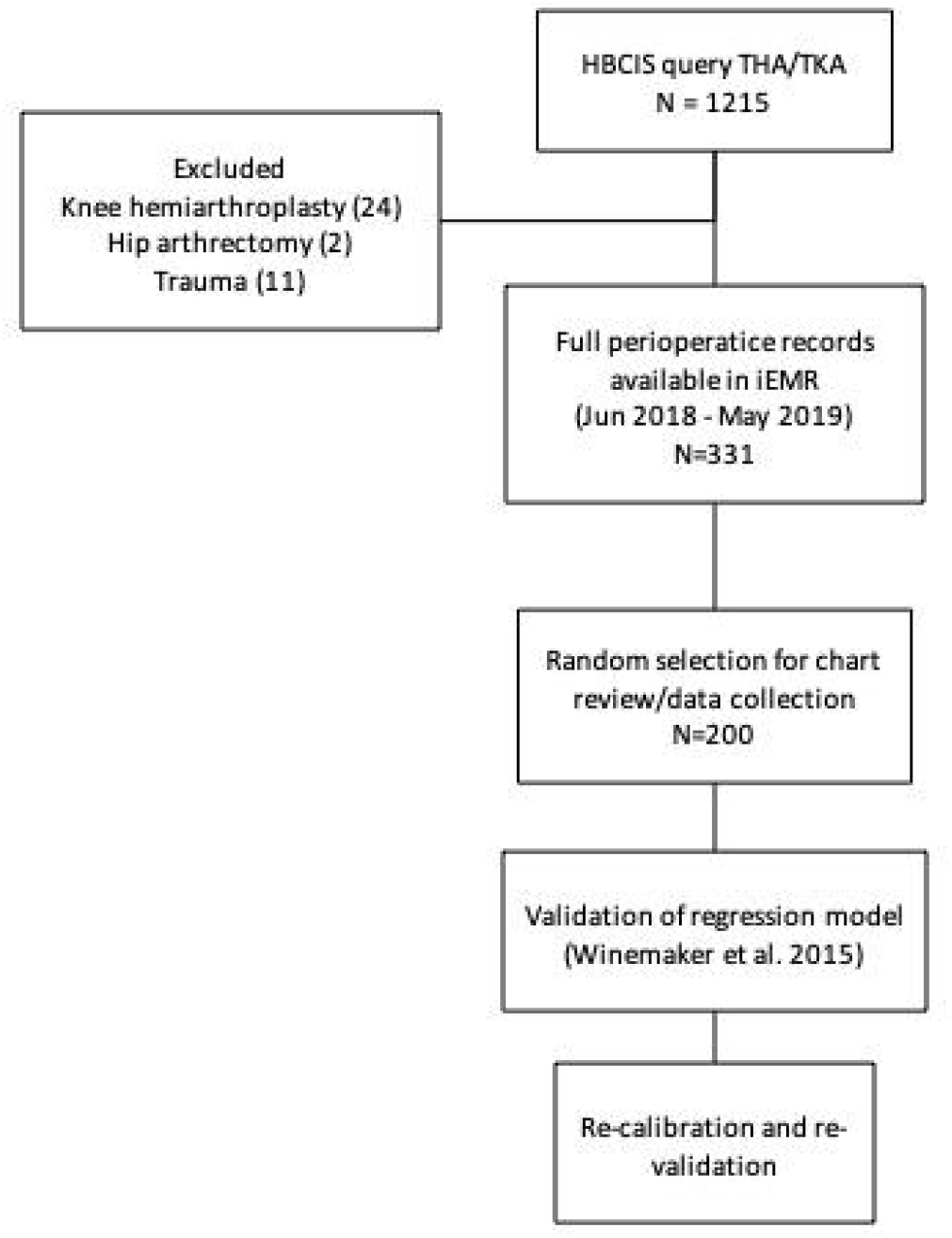
STROBE [22] flow diagram from initial hospital record query to model validation - 01-Jun-2018 to 04-Apr-2019. HBCIS, hospital based corporate information system; EMR, electronic medical record.

#### Treatment details

All patients electing to undergo total knee or hip arthroplasty for end-stage arthritis underwent standard multidisciplinary preoperative screening incorporating anaesthetic, orthopaedic, nursing and allied health review to ensure fitness for surgery. All eligible patients were subject to a standardised Enhanced Recovery After Surgery (ERAS) protocol involving the use of spinal anaesthesia and tranexamic acid, chemical and mechanical venous thromboembolism (VTE) prophylaxis and attempted mobilisation on the day of surgery. Fitness for discharge was assessed on a daily basis, defined as functional safety as determined by a qualified physiotherapist; and medically stable as determined by a medical practitioner. If the patient was identified by the physiotherapy team as requiring additional inpatient rehabilitation once declared medically stable, an orthogeriatric review was completed to enable referral to an external rehabilitation facility.

### Ethics and Governance

Ethical approval for the study was granted by the Metro South Human Research Ethics Committee (HREC/2019/QMS/57093). Data was retrieved by hospital staff from the electronic medical record and de-identified data was then distributed to the research team for further research, analysis and modelling.

### Multivariable Prediction Model

A literature search was conducted in Feb 2019 to identify potential models for identifying patients at risk of extended stay following in-patient total hip and knee arthroplasty.The three studies selected described predictive models based on commonly collected preoperative factors including demographics, comorbidities, mobility, use of opioid painkillers, and select psychosocial factors such as availability of community support.

The Winemaker model [18] was selected for assessment based on compatibility with the dataset available in the electronic medical record. This model used a generalised logistic regression model to fit to a three-level classification of LOS, with <3 days used as the reference level, 4 days and ≥5 days being the comparator levels. A stepwise elimination approach was applied to a starting predictor list of 102 factors using an α of 0.2. The original model was developed using SAS (v9.3, SAS Institute), R (v3.03, R Foundation, Austria) and SPSS (v20, IBM Corp, USA). Any internal validation process for the model was not described in the original paper.

### Development vs Validation

The primary outcome of this validation analysis was to identify the probability of an individual requiring a length of acute hospital stay following TJA equal to or greater than 5 days, or the probability of patients staying for the target 3 days or less. At present, current hospital processes rely on post operative factors to identify that a LOS of greater than 5 days is imminent, at which time a transfer to an extended stay rehabilitation facility is initiated. The end of surgery date and time (hh:mm), and discharge date and time were extracted by data linkage to a surgical data electronic system within the hospital to calculate LOS. For the purposes of model/tool assessment, the LOS was recategorised based on the threshold described in [18]. Other differences in setting and case-mix are summarised in **Table 1**.

**Table 1:** Comparison of methodology between development and validation analyses

|  | Development | Validation |
| --- | --- | --- |
| Setting | 10 orthopaedic surgeons at 1 high-volume hospital with academic affiliation in Ontario, Canada. | 14 orthopaedic surgeons at 1 high-volume public hospital in Queensland, Australia |
| Case eligibility/mix | Primary total hip or knee surgery and attended the preoperative clinic for medical history and assessment by the anesthesiology department Apr 2011 - Mar 2012 | Total hip or knee arthroplasty (primary or revision) with an electronic record available on the integrated medical record system; date of surgery Apr 2018 - Apr 2019 |
| Outcome | Length of stay categorised as 5 days or more (LOS not defined) | Length of stay categorised as 5 days or more (LOS defined as difference from date-time of surgery to date-time of discharge from ward) |

#### Predictors

The original publication listed a series of candidate variables in supplementary material, but did not provide explicit definitions. The predictors from the final model have been tabulated from the final published equations and inferences made from the publication text regarding reference categories, responses and definitions **(Table 2)**. A list of variables and outcomes used in the external validation set can be found in **Supplementary file A**.

**Table 2:** Predictors from final Winemaker model (retrieved from original supplementary material)

|  | Reference category | Definition (assumed) | Responses |
| --- | --- | --- | --- |
| Sex | Male | Sex identified at the time of surgery booking | Male;<br>Female |
| Joint | Hip | Articulation receiving arthroplasty in the current episode of care | Hip;<br>Knee |
| Age | 75+ | Biological age at the time of the procedure | <75;<br>≥75 |
| Smoking status | No | The patient's current smoking status at the time of preoperative assessment | No;<br>Yes |
| Diabetes, Oral treatment | No | Whether the patient was treating NIDDM with oral medications | No;<br>Yes |
| Diabetes, Insulin | No | Whether the patient was treating insulin-dependent diabetes with insulin | No;<br>Yes |
| Previous Stroke/TIA | No | Assessed by the anaesthetist at preoperative visit | No;<br>Yes |
| ASA | 1 | American Society of Anesthesiologists classification | 1;<br>2;<br>3; |
|  |  |  | 4 |
| Number cardiovascular comorbidities | NA | Assessed by the anaesthetist at preoperative visit | Count |
| Number of renal comorbidities | NA | Assessed by the anaesthetist at preoperative visit | Count |
| Other musculoskeletal comorbidities | No | Assessed by the anaesthetist at preoperative visit | No;<br>Yes |

NIDDM, non-insulin-dependent diabetes; TIA, transient ischaemic attack; ASA, American Society of Anesthesiologists

#### Risk Groups

The Winemaker model dichotomised two continuous predictors. Age was dichotomised to above and below 75 years of age at the time of surgery. American Society of Anaesthetist (ASA) classification is also a subjective rating of the patient’s physical status on an ordinal interval scale (1 to 4). This was dichotomized into 4 categories and ASA 1 left as the reference category, to create risk groups around physical status and comorbidity.

### Sample Size

Methods for estimating appropriate validation samples for external validation of binary outcome models [23] were used to calculate minimum sample sizes within acceptable margins of error. In brief, the sample size required for overall calibration (observed:expected ratio), calibration slope, c-statistic and standardised net benefit were estimated using the code provided **(Table 3)**. The key inputs were an original prevalence of 44% of patients staying >5 days (as reported in Winemaker) and a reported c-statistic of 0.73 for the 5-day model (retrieved from Supplementary material). The sample size retrieved was based on the availability of the data extraction team.

**Table 3:** Summary of sample size estimates for external validation

|  | Inputs | Sample Size | Events |
| --- | --- | --- | --- |
| Observed:Expected | Outcome proportion - 44% | 23 | 10 |
| Calibration Slope | Distribution<br>Mean: -1.3086<br>Variance: 17.1<br>Skewness: -1<br>Kurtosis: 4 | 1643 | 723 |
| C-statistic | Outcome proportion - 44%<br>C-Statistic: 0.73 | 392 | 173 |
| Standardised Net Benefit | Outcome proportion - 44%<br>Sensitivity: 0.85<br>Specificity: 0.86<br>Threshold Probability: 0.5 | 295 | 130 |

### Statistical Analysis

All patient datasets contained the necessary fields from the selected model (no missing data). The linear predictor (LP) was calculated on the validation dataset and predicted probabilities calculated using an inverse logit function (**Supplementary file B**). External validation of the Winemaker model was evaluated across the following domains as described in Riley et al 2021 [23]; All analyses were conducted using Stata (v17.1, Statacorp, USA).

#### Discrimination

A nonparametric Receiver Operating Curve analysis (roctab function) was applied to the predicted probabilities for the primary outcome. A brier score was calculated to assess the accuracy of probabilistic ratings generated by the model.

#### Model Calibration

Overall calibration was assessed by calculating the ratio between expected incidence and observed incidence of the primary outcome (>5 day LOS). Calibration-in-the-large was determined with a logistic regression applied to the predicted probabilities with offset applied (calculated LP) [24]. Calibration slope was assessed by reapplying the logistic regression without an offset. A calibration plot was generated using the pmcalplot function [25].

#### Model Update

External validation metrics for the original model equation were stored **(Supplementary file B)**. The model equation was updated as per [24,26] to take into account a corrected intercept and to shrink the model coefficients to account for model optimism during development. The external validation functions were then rerun and the outputs reported.

#### Net Benefit

Net benefit was calculated on the updated model using methods described by Riley et al 2021 [23], and interpreted according to guidance outlined in Vickers et al., 2019 [27]. Threshold probability of 0.5 was selected as clinically acceptable for the ratio of correct to unnecessary earlier initiations of postoperative planning for the use of community support services or referrals to inpatient rehabilitation. A 1:1 ratio was considered acceptable as the harms of the proposed intervention are largely administrative.

## Results

### Patient characteristics

A random selection of 200 patients was extracted for analysis, comprising a majority female and obese sample **(Table 4)**. The majority of TJAs (89%) were performed for osteoarthritis, with 83 hip procedures and 117 knee procedures. The majority of patients (70%) presented with at least one cardiovascular comorbidity (hypertension or atherosclerosis), with endocrine (47.5%) and gastrointestinal diagnoses (43%) also commonly noted.

**Table 4:** Comparison between the development sample (Winemaker) and the validation sample retrieved for the present analysis

|  | Development | Validation |
| --- | --- | --- |
| Prevalence ($\geq 5$ days LOS) | 44 | 24 |
| Sample | 1459 | 200 |
| Male (%) | 42 | 40.5 |
| Age (median) | 67 (52 - 82) | 70 (62.5 - 75) |
| Age $\geq 75$ | 26.7 | 25.5 |
| Knee (%) | 61.7 | 58.5 |
| Smoking (%) | 11.6 | 9 |
| Diabetes (%) | 29.5 | 16.5 |
| Diabetes Oral Medication (%) | Not reported | 12.5 |
| Diabetes Insulin (%) | Not reported | 4 |
| Previous Stroke/TIA (%) | 7.5 | 5 |
| ASA (%) | 1 - 0.8<br>2 - 23.6<br>3 - 62<br>4 - 13.6 | 1 - 1.0<br>2 - 55.6<br>3 - 42.9<br>4 - 0.5 |
| Number of cardiovascular comorbidities | Not reported | 0 - 31<br>1 - 51.5<br>2 - 11.5<br>3 - 3<br>4 - 1<br>5 - 2 |
| Number of renal comorbidities (median) | Not reported | 0 - 87.5<br>1 - 12.5 |
| Other musculoskeletal comorbidity (%) | 14.6 | 19.5 |

TIA, Transient ischaemic attack; ASA, American Society of Anaesthesiologists classification

The LOS ranged from 1.8 to 20 days at a median of 3.8 days (IQR 3 - 5), with 60% of patients discharged within 3 days of surgery, 16% of patients discharged at 4 days, and 24% at 5 days or greater. Of the patients in the ‘at risk’ category, the median LOS was 6 days (IQR 4-8). The majority (93%) of patients were discharged home with community support defined as in the care of relatives, spouses or carers; 5% required admission to inpatient rehabilitation, and 2% were discharged home to recover independently.

### Data coverage

The Winemaker model was validated against 200 patients, with sample characteristics summarised in **Table 4**. A breakdown of sample characteristics was not fully reported in the original paper.

#### External Validation - Original

The area under the ROC (AUROC) was estimated at 0.64 (95%CI 0.56 - 0.72). The ratio of the expected frequency of cases (74.1%) to have LOS >5 days to the observed frequency (31.5%) was 2.35. The Brier score was 0.39 with Spiegelhalter’s Z-statistic of 16.9 (P < 0.001). Mean calibration was determined via the calibration intercept (-3.59, SE 0.17, 95%CI -3.92 to -3.24). Calibration of the Winemaker model demonstrated gross overestimation (calibration slope 0.52) of the risk of >5 day LOS in the validation sample **(Figure 2)**, with large variances in the estimates at each percentile and a non-linear curve from lowest to highest risk subgroups.

**Figure 2:**
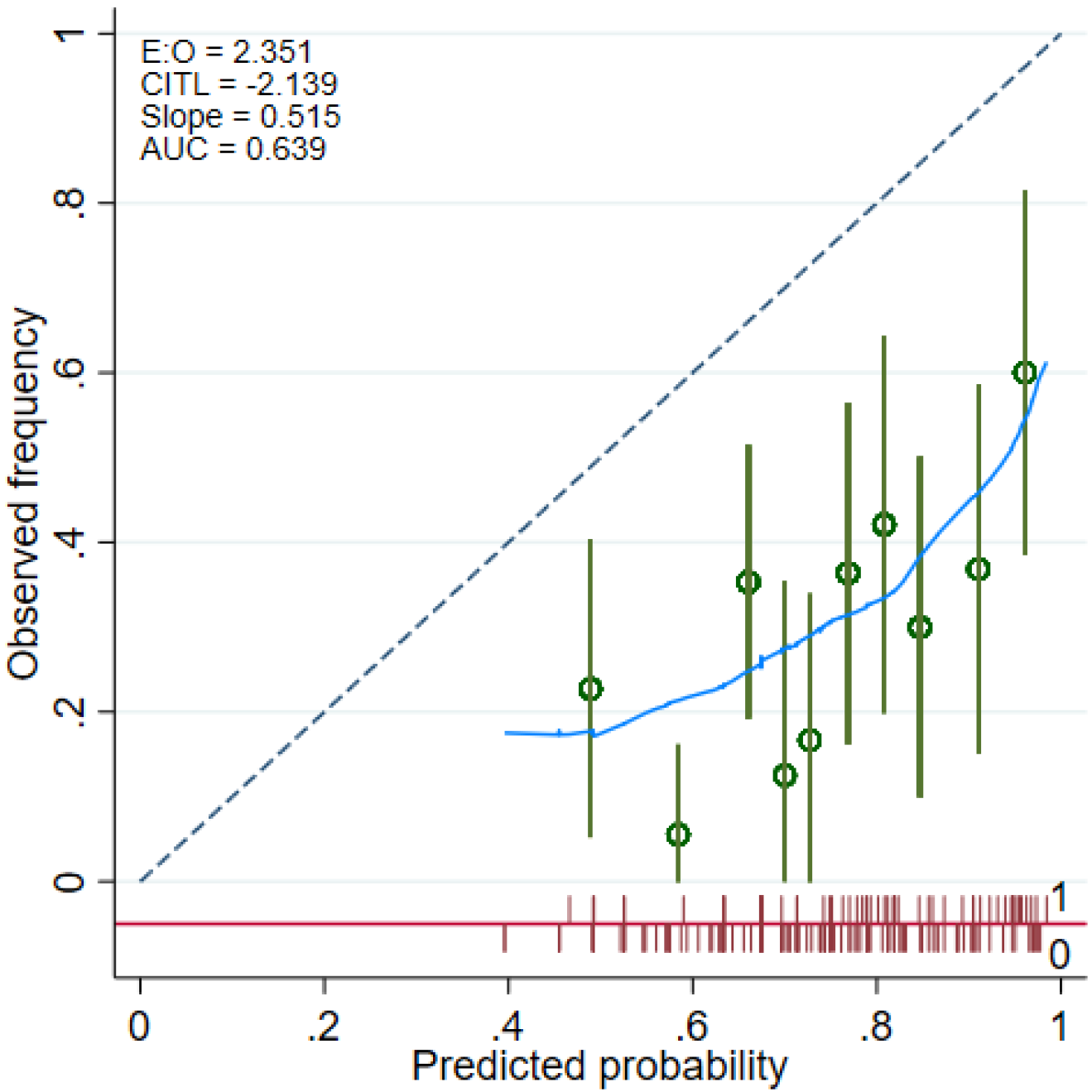
Calibration curve of the Winemaker model (Original) applied to the validation dataset

#### External Validation - Model Recalibration

The model intercept was updated to the validation data and the model coefficients rescaled. The area under the ROC (AUROC) was estimated at 0.69 (95%CI 0.61 - 0.77). The Brier score was 0.197 with Spiegelhalter’s Z-statistic of 0.0497 (P = 0.481). Mean calibration was determined via the calibration intercept (0.48, SE 0.16, 95%CI 0.16 to 0.80). Calibration of the *updated* model demonstrated was improved for the risk of >5 day LOS in the validation sample **(Figure 3)**, but with large variances in the estimates at each percentile.

**Figure 3:**
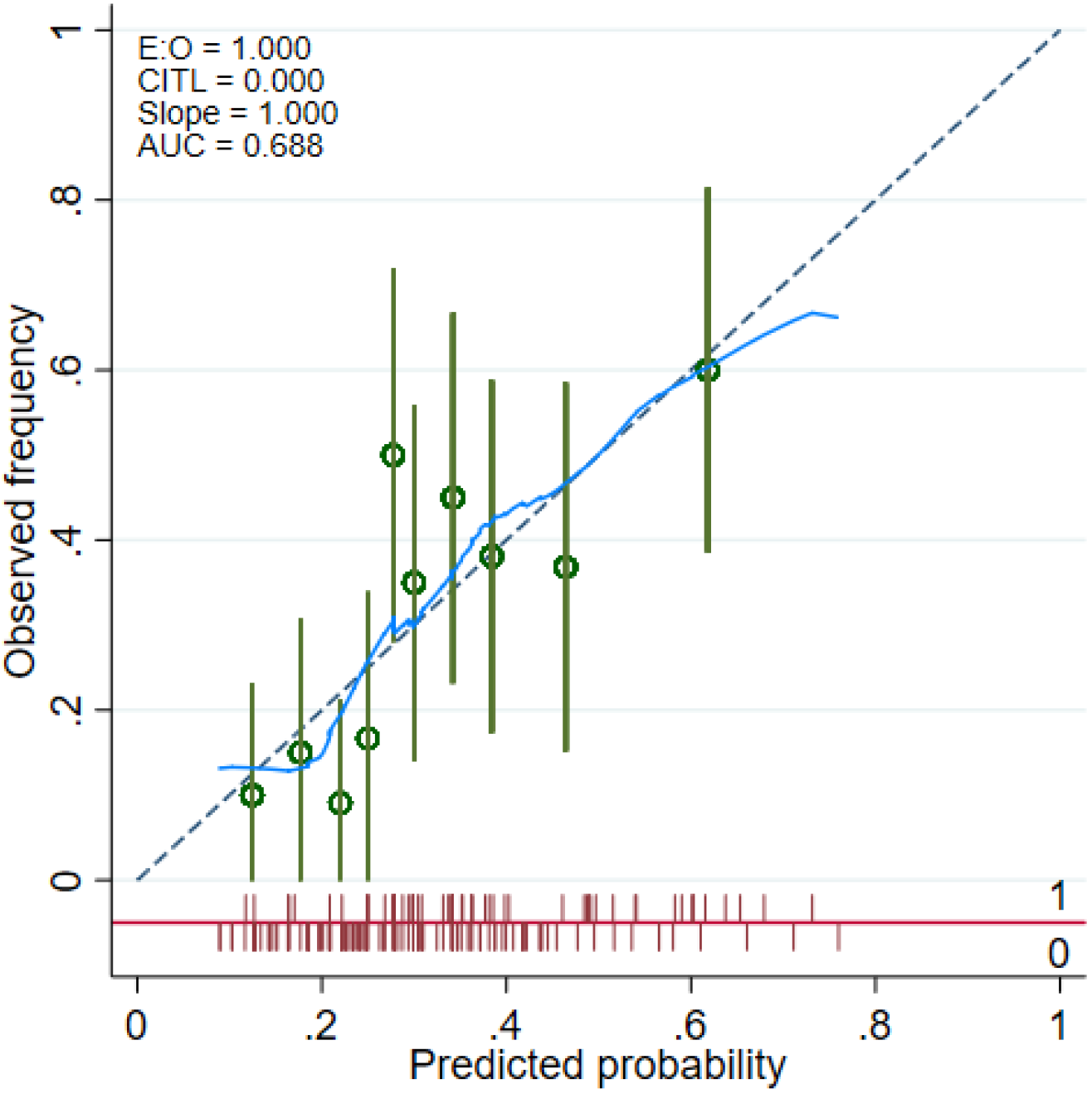
Calibration curve of the Winemaker model (Updated) applied to the validation dataset

#### Clinical Utility

The updated model showed limited clinical utility above usual care at the a-priori probability threshold (0.5), with a net benefit of approximately 0.03; representing 3 patients per hundred correctly identified for earlier initiation of postoperative planning **(Figure 4)**. At lower risk thresholds (∼0.25), the model demonstrated a net benefit superior to treating all; with approximately 15 patients per hundred correctly identified prior to surgery.

**Figure 4:**
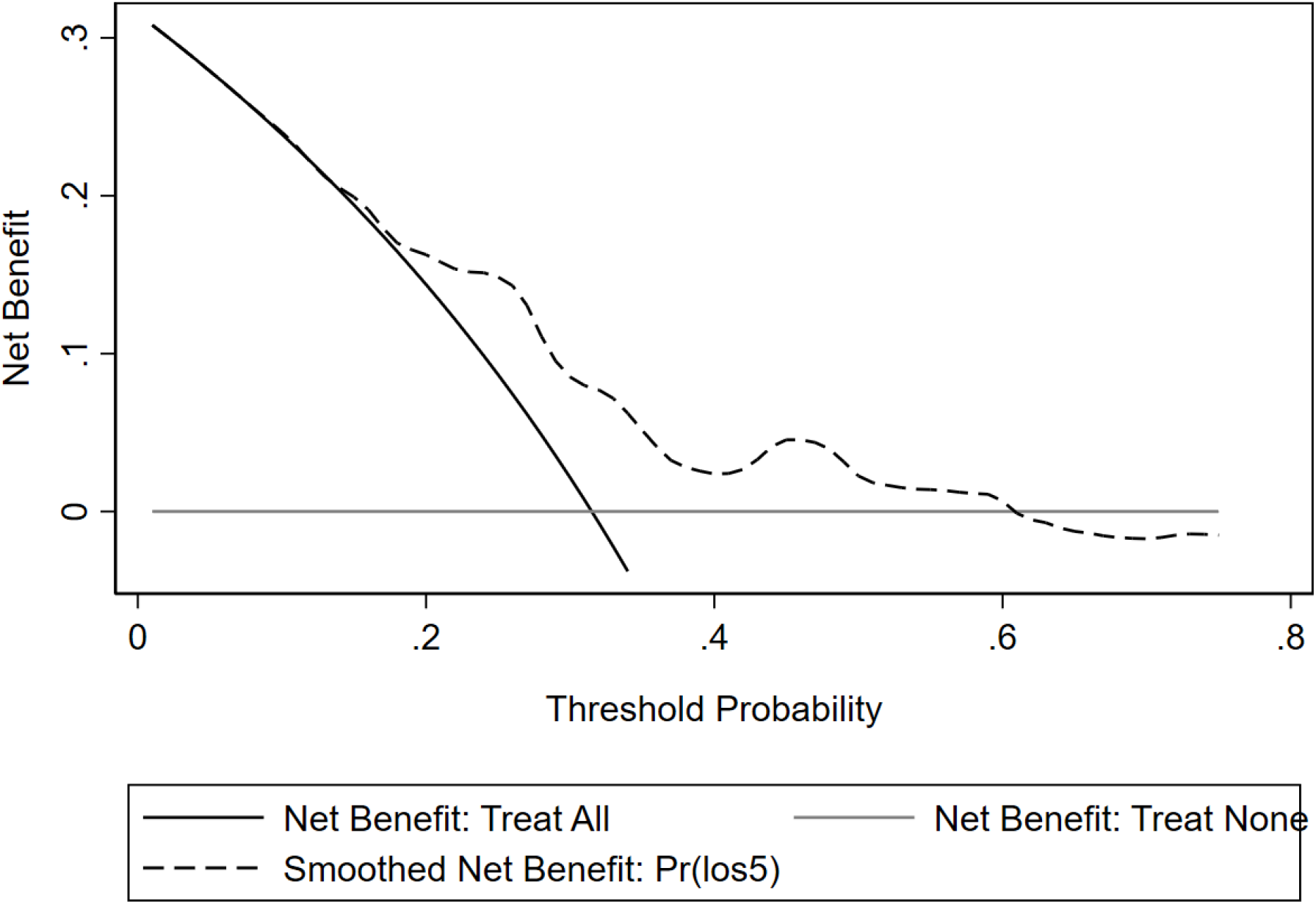
Decision curve analysis of the adjusted model

## Discussion

### Summary

Predictive model development for hospital decision-making is dependent on quality data linkage, and sufficient volume of patients to reach adequate sample sizes for development and internal validation [28]. The present study attempted to externally validate a predictive model for classifying patients at risk of an extended hospital stay after TJA to determine the clinical relevance of such tools across a broad range of settings. In validating the model to an independent sample, the transportability and clinical utility are able to be determined to ensure such models are applied accurately. The present validation demonstrated that when applied to a new population the selected model [18] grossly overestimated the risk of extended LOS and as a result could have inaccurately directed clinical care and may not be suitable for introduction into clinical practice within the chosen setting. The unadjusted model was poorly calibrated to the data and while the adjusted model displayed improvements, it remained poorly calibrated in low to medium risk patients. These patterns of miscalibration have previously been attributed to under-specification within the model, that is key risk factors have been left unaccounted [12,29]. In addition, the adjusted model offers limited benefits above usual care.

### Model Calibration

The selected model displayed miscalibration in predicting risk, particularly in patients with low to medium risk profiles. The lack of calibration can be explained by a number of factors. Firstly, the model development involved stepwise variable selection. Stepwise selection (based on significant relationships) and other forms of automated variable selection are limited in this context, with potential for mislabeling of important variables [30] under the false assumption that only important predictive variables display significance [31]. This may have contributed to other important factors being omitted from the model. Secondly, no internal validation process was implemented, which may have produced overly optimistic regression coefficients. Thirdly, a number of the variables included in the original model are open to subjective interpretation, both at the clinical level (data input) and by the observer reviewing case notes (data extraction). Lastly, perioperative management practices regarding LOS have evolved greatly since the period the model was originally developed (∼2012). Clinical and administration culture, attitudes and policies towards shorter stay have evolved over this time.

### Clinical Utility

The clinical utility (0.03 at threshold probability of 0.5) for the selected model was lower compared to other published models (eg. machine learning) [32], with net benefit of approximately 0.17 - 0.25 at the same probability threshold. Overall, the net benefits observed in this study were modest, with little net benefit (approximately 3 patients per hundred correctly flagged for ‘intervention’ prior to surgery) associated with the chosen probability threshold. However, net benefit of a more conservative threshold cutoff (0.25) was superior to the ‘treat all’ scenario, correctly identifying 15 at-risk patients per hundred prior to surgery. The ability to reduce LoS in key patient subgroups depends on the structure and cost of i) implementing the model adjusted for the local setting into clinical practice, and ii) the selected intervention(s) to act on the predictions. A low-cost implementation and intervention has the potential to free up ward bed-days and reduce average LOS if other cost drivers (e.g. hospital readmission) are not impacted. A relatively low cost implementation may involve a digital application (e.g. [33]) that can be integrated into an electronic medical record. Further consideration of financial and other costs are required to justify implementing the model in its current form.

### Implications and Future Directions

Overall, the selected model performance poses considerable challenges to implementing into clinical practice and guiding suitable interventions to reduce LOS in the local setting. The implications of this finding are that the use of published models and tools without additional validation within the intended use setting may introduce miscalibration for risk prediction. For example the RAPT is a popular tool for discharge planning after lower limb arthroplasty, but has yet to be externally validated for LOS. However, examples of its routine use in predicting LOS in arthroplasty settings outside of its development [12] and attempts at external validation [34] have been reported [35]. Clinical centres using tools in this way should consider a validation process to identify the need for adjusting the model to the local population, or halting use of the model if it is found to be unsuitable. Indeed, even suitable models implemented over long periods of time are at risk of calibration drift [36] and may need to be updated regularly [37].

With respect to the underlying motivation for this study, the health service still seeks an appropriate model to guide intervention selection and perioperative management redesign. Selecting an appropriate model for implementation remains challenging, although there are numerous published models in the literature [14], the quality of medical prognostic models are generally low [38] with bias introduced through analytical decisions, internal validation (correcting for model optimism) poorly reported and even fewer models with demonstrated external validation (transportability). Possible candidate models may be specific for total knee [39], total hip [40] or combined [41]. A review of the candidate variables used across all the available models display a broad spectrum of patient, pathology, and administrative characteristics that have been used in a variety of statistical, machine-learning and other model types. More recent work has identified other potential variables that could be included in model development that may build on previous work. For example, patients with fewer comorbidities, patient-reported psychosocial risk factors or functional deficits are crucial for differentiating between those who are likely to leave hospital within the target time frame, and those who stay longer [12,42]. Others have reported other administrative aspects such as day of the week that surgery is performed may have a bearing on LOS [43]. In the context of data constraints, predictive modelling may not require individual patient-level precision in order to be clinically useful [44], and such tools do not replace the patient-level decision making which takes place prior to surgery. Therefore, selection of the ideal predictive model for clinical adaptation requires a detailed consideration of how it will integrate into the clinical decision-making process. Further work is required to better identify candidate models and predictors to develop prognostic models in local settings or within larger clinical networks.

### Limitations

The findings of the present study to validate a published model in a new hospital setting should be interpreted in light of its limitations. Firstly, the overall data pool was restricted to a single year due to changeover in electronic hospital systems, limiting the sample size available for validation. Secondly, while hospital-level data is unlikely to be detailed enough (and may not be necessary) to capture all relevant factors affecting a patient’s LOS, classification of certain comorbidities may impact their predictive accuracy. For example, cardiovascular comorbidities include conditions which are relatively simple to manage (eg. hypertension), as well as those likely to require additional procedures to manage risk during surgery, such as atrial fibrillation. The subjective nature of both data input and the point of care and the interpretation of notes by chart review may be mitigated in future by standardised capture and reporting methods. Thirdly, the sample size calculation indicated a much larger sample than was available for this analysis is required to describe the calibration slope. While it is acknowledged some miscalibration could be explained by the reduced sample, it may not fully explain the miscalibration observed and caution should still be applied to implementing the model in its current form into the target setting.

## Conclusion

This study reports an external validation of a regression model intended to identify patients at risk of an extended hospital stay after total hip or knee arthroplasty. To our knowledge, this is the first attempt at applying the model to a different clinical setting. Transportability of this model to a smaller dataset was hampered by differences in incidence of the primary outcome, leading to a substantial overestimation of risk. Distribution of risk factors could not be compared as they were not reported clearly in the original dataset. Estimates of clinical benefit strongly suggest clinical decision-making based on these risk factors would be of only modest benefit to patients. It is recommended that attempts to improve planning of postoperative care first focus on identifying factors that differentiate between individuals classified at ‘moderate risk’.

## Supporting information

Supplementary file A: Variable and outcome definitions

Supplementary file B: Statistical analysis output

TRIPOD Checklist

## Data Availability

Data may be available from the corresponding author upon reasonable request.

## Acknowledgements

The authors thank the nursing and allied health staff at the Queen Elizabeth II Jubilee Hospital, Brisbane, Qld Australia and the data analysis staff at EBMA Analytics, Sydney, NSW Australia for support and assistance with this study. The authors also acknowledge the assistance of Kerry Baldwin (QEII Physiotherapy) and Maha Jegatheesan (QEII Orthopaedics) for their contributions to chart review, and Thomas Cooper (EBMA) for his contributions to the analysis code. The authors also gratefully acknowledge Dr Lorenzo Calabro for his valuable feedback on the manuscript.

