## Supplementary file A: Variable and outcome definitions for "Applying models of care for total hip and knee arthroplasty: External validation of a published predictive model to identify extended stay risk prior to lower-limb arthroplasty"

### Supplementary material A: Variable and outcome definitions

**Age**
*Age at time of surgery, in years*
Age 75 years or older

**Height**
*In metres, to calculate BMI*

**Weight**
*In kg, to calculate BMI*

**Gender**
Male;
Female;
Not specified

**Operation Type description**
Elective;
Emergency;
Not specified

**Bilateral status**
Unilateral;
Simultaneous;
Sequential

**Diagnosis**
*Primary diagnosis related to procedure*
Osteoarthritis;
Rheumatoid Arthritis;
Psoriatic Arthritis;
Septic Arthritis

**Joint**
Knee;
Hip

**ASA score**
1;
2;
3;
4

**Date of surgery**
Separated ‘In OR At’ variable from dd mmm yyyy HH:MM:ss AM/PM’ into ‘Date of surgery’ and ‘Time of surgery (in)’

Converted the date component to dd-mmm-yyyy

**Time of surgery (out)**
Separated ‘Out OR At’ variable from dd mmm yyyy HH:MM:ss AM/PM and converted the time component into HH:MM:ss 24 hour time

**Date of discharge**
Separated ‘Discharge DateTime’ variable from dd mmm yyyy HH:MM:ss AM/PM’ into ‘Date of discharge’ and ‘Time of discharge’
Converted the date component into dd-mmm-yyyy

**Time of discharge**
Converted from HH:MM:ss AM/PM into HH:MM:ss 24hour time.

**Length of Stay**
Date and time of surgery to date and time of discharge (in days)

**Discharge destination**
Home;
Home with community support;
Inpatient rehab;
Slow stream rehab;
Aged care

#### Comorbidities:

**Cardiovascular**
Yes; No
*Common inclusions: Hypertension; dysrhythmia; valvular disease; congestive heart failure; decreased left ventricular function; peripheral vascular disease; ischaemic heart disease; previous MI; stable angina; unstable angina; stent; coronary artery bypass grafting*

**Cardiovascular (specify)**
*Each cardiovascular comorbidity listed, semicolon delimited for calculation of ‘number of cardiovascular comorbidities’*

**Diabetes**
Yes - oral medication;
Yes - Insulin;
No

**Previous stroke/TIA**
Yes; No

**Renal**
Yes; No
*Common inclusions: Renal insufficiency; renal failure; hemodialysis; shunt/fistula; peritoneal dialysis*

**Musculoskeletal (other than index pathology)**
Yes; No
*Common inclusions: Back pain diagnosis; chronic pain; fibromyalgia; ankylosing spondylitis; spina bifida*

**Others**
S*moking status if listed.*
