## Supplementary file B: Statistical analysis output for "Applying models of care for total hip and knee arthroplasty: External validation of a published predictive model to identify extended stay risk prior to lower-limb arthroplasty"

**Supplementary material B**: Statistical analysis output

```
                                                                     Statistics/Data
                                                                  analysis

                                          User: Pub_Extended LOS ReAnalysis_CB041Jan22 Fourth Pass
                                                Project: Pub_Extended LOS ReAnalysis_CB041Jan22

 1 . clear

 2 . do "C:\Users\cscho\AppData\Local\Temp\STD1a38_000000.tmp"

 3 . clear

 4 . *Set working directory
 5 . *Example given below, replace with the path to your working folder
 6 . cd "G:\My Drive\EBMA\Client Drive\QEII Jubilee\EBMA Working\Publications\Predicting extended stay\Update2022"
   G:\My Drive\EBMA\Client Drive\QEII Jubilee\EBMA Working\Publications\Predicting extended stay\Update2022

 7 .
 8 . *Load validation dataset
 9 . import delimited using "MasterWinemakerv2.csv", stringcols(9
   19) (encoding automatically selected: UTF-8)
   (19 vars, 230 obs)

10 .
   end of do-file

11 . do "C:\Users\cscho\AppData\Local\Temp\STD1a38_000000.tmp"

12 . *rename model inputs
13 . rename diabetesmedicationoral dmoral

14 . rename diabetesmedicationinsulin dminsulin

15 . rename previousstroketia stroketia

16 . rename musculoskeletalother      msk

17 . rename lengthofstaydostodischarge los

18 .
19 .
20 . * encode categorical variables
21 . label define sexcode 0 "female" 1 "male"

22 . encode gender, gen(sexcode)

23 . label define jointcode 0 "knee" 1 "hip"

24 . encode joint, gen(jointcode)

25 . label define dmcode 0 "No" 1 "Yes"

26 . encode dmoral, gen(dmoralcode)

27 . label define dminsulincode 0 "No" 1 "Yes"

28 . encode dminsulin, gen(dminsulincode)

29 . label define stroketiacode 0 "No" 1 "Yes"
```

```
30 . encode stroketia, gen(stroketiacode)

31 . label define mskcode 0 "No" 1 "Yes"

32 . encode msk, gen(mskcode)

33 . label define smokecode 0 "No" 1 "Yes"

34 . encode smoking, gen(smokecode)

35 . label define asa3code 0 "No" 1 "Yes"

36 . encode asa3, gen(asa3code)

37 . label define asa4code 0 "No" 1 "Yes"

38 . encode asa4, gen(asa4code)

39 .
40 . encode agecat, gen(agecatcode)

41 . encode asascore, gen(asacode)

42 .
43 . ***Create binary outcome variables*****
44 . generate los3 = 0

45 . replace los3 = 1 if los
   <= 3 (106 real changes
   made)

46 .
47 . generate los4 = 0

48 . replace los4 = 1 if
   inlist(los,4) (48 real changes
   made)

49 .
50 . generate los5 = 0

51 . replace los5 = 1 if los
   >= 5 (76 real changes made)

52 .
   end of do-file

53 . do "C:\Users\cscho\AppData\Local\Temp\STD1a38_000000.tmp"

54 . *Check missing pattern of key variables
55 . misstable summarize
```

| | | | | Obs<. | | |
|---|---|---|---|---|---|---|
| Variable | Obs=. | Obs>. | Obs<. | Unique values | Min | Max |
| dmoralcode | **30** | | **200** | 2 | 1 | 2 |
| dminsulinc~e | **30** | | **200** | 2 | 0 | 1 |
| stroketiac~e | **30** | | **200** | 3 | 0 | 2 |
| mskcode | **30** | | **200** | 3 | 0 | 2 |
| asacode | **32** | | **198** | 4 | 1 | 4 |

```
56 . misstable patterns dmoralcode dminsulincode stroketiacode mskcode los5

      Missing-value
        patterns (1 means
        complete)

              |   Pattern
      Percent |  1  2  3  4
     ---------+-------------
         87%  |  1  1  1  1
              |
         13   |  0  0  0  0
     ---------+-------------
        100%  |

     Variables are (1) dminsulincode (2) dmoralcode (3) mskcode (4) stroketiacode

57 .
58 . *Insert Multiple Imputation here if needed
59 .
60 .
61 . *Calculate the linear predictor and predicted probability for each individual in the
62 . *validation dataset. Calculate the mean and standard deviation of the linear predictor.
63 .
64 . // REPLACED ORIGINAL EQUATIONS MANUALLY FROM THE PAPER (JE Updated Mar-Apr 2022)
65 . gen lin_pred4= -0.5115 + (-0.7358*sexcode) + (0.7148*jointcode) + (0.4938*agecatcode) + (-0.8565*smokecode) + (0.7181
   > *dminsulincode) + (0.3099*stroketiacode) + (-0.2968*[asacode==3]) + (0.3860*[asacode==4]) + (0.1183*nocvd) + (0.5161*
   > ode)
   (30 missing values generated)

66 .
67 . gen lin_pred5= -1.3086 + (-0.5551*sexcode) + (0.9676*jointcode) + (1.4177*agecatcode) + (-0.7145*smokecode) + (0.2438
   > dminsulincode) + (0.0834*stroketiacode) + (0.5437*[asacode==3]) + (1.1077*[asacode==4]) + (0.2770*nocvd) + (1.1745*no
   > e)
   (30 missing values generated)

68 .
   end of do-file

69 . do "C:\Users\cscho\AppData\Local\Temp\STD1a38_000000.tmp"

70 . // EXPANDED THE PROB CALCS FOR 4 AND 5 TO INCLUDE THE EXTRA EXP()
71 . gen prob4=
   exp(lin_pred4)/(1+exp(lin_pred4)+exp(lin_pred5)) (30
   missing values generated)

72 .
73 . gen prob5 =
   exp(lin_pred5)/(1+exp(lin_pred4)+exp(lin_pred5)) (30 missing
   values generated)

74 .
75 . gen prob3 =
   1/(1+exp(lin_pred4)+exp(lin_pred5)) (30 missing
   values generated)

76 .
77 . // Calculate the C-statistic to evaluate the discrimination of the model.
78 .
```

```
79 . *Discrimination
80 . roctab los3 prob3
```

| Obs | ROC area | Std. err. | Asymptotic normal [95% conf. interval] | |
|---|---|---|---|---|
| 200 | 0.6570 | 0.0388 | 0.58092 | 0.73317 |

```
81 .
82 . roctab los5 prob5
```

| Obs | ROC area | Std. err. | Asymptotic normal [95% conf. interval] | |
|---|---|---|---|---|
| 200 | 0.6387 | 0.0423 | 0.55579 | 0.72158 |

```
83 .
84 . *Alternative (but no CI)
85 . brier los3 prob3

Mean probability of outcome    0.4750
                 of forecast 0.0852

   Correlation                    0.2367
   ROC area                       0.6570 p = 0.0001

   Brier score                    0.3886
   Spiegelhalter's z-statistic 23.0850 p = 0.0000
   Sanders-modified Brier score
   0.3874 Sanders resolution   0.2189
   Outcome index variance         0.2494
   Murphy resolution              0.0305
   Reliability-in-the-small       0.1686
   Forecast variance              0.0069
   Excess forecast variance       0.0065
   Minimum forecast variance      0.0004
   Reliability-in-the-large       0.1520
   2*Forecast-Outcome-Covar       0.0197

86 .
87 . brier los5 prob5

Mean probability of outcome    0.3150
                 of forecast   0.7406

   Correlation                    0.2213
   ROC area                       0.6387   p = 0.0008

   Brier score                    0.3879
   Spiegelhalter's z-statistic   16.1925   p = 0.0000
   Sanders-modified Brier score   0.3888
   Sanders resolution             0.1955
   Outcome index variance         0.2158
   Murphy resolution              0.0203
   Reliability-in-the-small       0.1933
   Forecast variance              0.0200
   Excess forecast variance       0.0190
   Minimum forecast variance      0.0010
   Reliability-in-the-large       0.1812
   2*Forecast-Outcome-Covar       0.0291
```

```
 88 .
    end of do-file

 89 . do "C:\Users\cscho\AppData\Local\Temp\STD1a38_000000.tmp"

 90 . For calibration, first calculate measures of overall calibration i.e. E/O and
    command For not defined by For.ado
    r(199);

    end of do-file

    r(199);

 91 . do "C:\Users\cscho\AppData\Local\Temp\STD1a38_000000.tmp"

 92 . *Expected/observed
 93 . summ prob5
```

| Variable | Obs | Mean | Std. dev. | Min | Max |
|---|---|---|---|---|---|
| prob5 | **200** | **.7406444** | **.1416446** | **.3957578** | **.984455** |

```
 94 . summ los5
```

| Variable | Obs | Mean | Std. dev. | Min | Max |
|---|---|---|---|---|---|
| los5 | **230** | **.3304348** | **.4713956** | **0** | **1** |

```
 95 . di "E/O = "0.7386/0.33
    E/O = 2.2381818

 96 .
 97 . *Calibration-in-the-large
 98 . logistic los5, offset(lin_pred5) coef

Logistic regression                                   Number of obs = 200
                                                      Wald chi2(0)  =    .
Log likelihood = -120.48978                           Prob > chi2   =    .
```

| los5 | Coefficient | Std. err. | z | P>\|z\| | [95% conf. | interval] |
|---|---|---|---|---|---|---|
| _cons | **-3.589179** | **.1737921** | **-20.65** | **0.000** | **-3.929805** | **-3.248553** |
| lin_pred5 | **1** | (offset) | | | | |

```
 99 .
100 . *Calibration plot
101 . /* NB: using the 'keep' option will store the expected, observed and risk group
    >         variables used for the plot */
102 . pmcalplot prob5 los5, ci keep name(calibration_plot, replace) xtitle("Predicted probability", size(medsmall)) ytitle(
    > ize(medsmall)) legend(off)
    Binary option selected: Calibration plot for logistic prediction model displaying...

103 .
```

```
104 . graph save "calibration_plot" "G:\My Drive\EBMA\Client Drive\QEII Jubilee\EBMA Working\Publications\Predicting extend
    > bration_plot - Initialv2.gph", replace
    file G:\My Drive\EBMA\Client Drive\QEII Jubilee\EBMA Working\Publications\Predicting extended stay\Update2022\calibrati
    >  saved

105 .
106 . graph export "G:\My Drive\EBMA\Client Drive\QEII Jubilee\EBMA Working\Publications\Predicting extended stay\Update202
    > itialv2.png", as(png) name("calibration_plot") replace
    file G:\My Drive\EBMA\Client Drive\QEII Jubilee\EBMA Working\Publications\Predicting extended stay\Update2022\calibrati
        Initialv2.png saved as PNG format

107 .
    end of do-file

108 . do "C:\Users\cscho\AppData\Local\Temp\STD1a38_000000.tmp"

109 . graph close

110 .
    end of do-file

111 . do "C:\Users\cscho\AppData\Local\Temp\STD1a38_000000.tmp"

112 . * check the expected and observed probabilities for the 10th risk group
113 . su obs_pmcalplot exp_pmcalplot if groups_pmcalplot==10

        Variable |        Obs        Mean    Std. dev.       Min        Max
    -------------+---------------------------------------------------------
    obs_pmcalp~t |         20          .6           0         .6         .6
    exp_pmcalp~t |         20    .9607756           0   .9607756   .9607756

114 .
115 .
116 . ***Save data file***
117 . save "WinemakerValidv2.dta",
    replace file WinemakerValidv2.dta
    saved

118 .
    end of do-file

119 . do "C:\Users\cscho\AppData\Local\Temp\STD1a38_000000.tmp"

120 . *First need the linear predictor without the intercept
121 . gen lp_noint=lin_pred5 +
    1.30086 (30 missing values
    generated)

122 .
123 . /*Fit logistic model using lp_noint as the only predictor, this recalculates
    >         the intercept and scales the coefficients as needed in the validation cohort */
124 . logistic los5 lp_noint, coef

    Logistic regression                                     Number of obs =    200
                                                            LR chi2(1)    =  18.11
                                                            Prob > chi2   = 0.0000
    Log likelihood = -115.55546                             Pseudo R2     = 0.0727

    ------------------------------------------------------------------------------
            los5 | Coefficient  Std. err.      z    P>|z|     [95% conf. interval]
    -------------+----------------------------------------------------------------
        lp_noint |   .5433092   .1354295     4.01   0.000     .2778722    .8087462
           _cons |  -2.976091   .5841684    -5.09   0.000     -4.12104   -1.831142
    ------------------------------------------------------------------------------
```

```
125 .
126 . *Calculate the LP & predicted probabilites for the recalibrated model
127 . predict lin_pred5b, xb
    (30 missing values generated)

128 . predict prob5b, pr
    (30 missing values generated)

129 .
130 . *Calibration plot
131 . /* NB: using the 'keep' option will store the expected, observed and risk group
    >         variables used for the plot */
132 . pmcalplot prob5b los5, ci keep name(calibration_plot, replace) xtitle("Predicted probability", size(medsmall)) ytitle
    > size(medsmall)) legend(off)
    Binary option selected: Calibration plot for logistic prediction model displaying...
    Variable with name 'obs_pmcalplot' already exists, pmcalplot cannot generate required variables
    Variable with name 'exp_pmcalplot' already exists, pmcalplot cannot generate required variables
    Variable with name 'groups_pmcalplot' already exists, pmcalplot cannot generate required variables

133 .
134 . graph save "calibration_plot" "G:\My Drive\EBMA\Client Drive\QEII Jubilee\EBMA Working\Publications\Predicting extend
    > bration_plot - Model Updatedv2.gph", replace
    file G:\My Drive\EBMA\Client Drive\QEII Jubilee\EBMA Working\Publications\Predicting extended stay\Update2022\calibrati
    > v2.gph saved

135 .
136 . graph export "G:\My Drive\EBMA\Client Drive\QEII Jubilee\EBMA Working\Publications\Predicting extended stay\Update202
    > del Updatedv2.png", as(png) name("calibration_plot") replace
    file G:\My Drive\EBMA\Client Drive\QEII Jubilee\EBMA Working\Publications\Predicting extended stay\Update2022\calibrati
        Updatedv2.png saved as PNG format

137 .
138 . graph close

139 .
    end of do-file

140 . do "C:\Users\cscho\AppData\Local\Temp\STD1a38_000000.tmp"

141 . *First need the linear predictor without the intercept
142 . gen lp_noint=lin_pred5 +
    1.30086 variable lp_noint
    already defined r(110);

    end of do-file

    r(110);

143 . do "C:\Users\cscho\AppData\Local\Temp\STD1a38_000000.tmp"

144 . logistic los5 lp_noint, coef

    Logistic regression                                     Number of obs =     200
                                                            LR chi2(1)     =  18.11
                                                            Prob > chi2    = 0.0000
    Log likelihood = -115.55546                             Pseudo R2      = 0.0727
```

| los5 | Coefficient | Std. err. | z | P>\|z\| | [95% conf. | interval] |
|---|---|---|---|---|---|---|
| lp_noint | .5433092 | .1354295 | 4.01 | 0.000 | .2778722 | .8087462 |
| _cons | -2.976091 | .5841684 | -5.09 | 0.000 | -4.12104 | -1.831142 |

```
145 .
146 . *Calculate the LP & predicted probabilites for the recalibrated model
147 . predict lin_pred5b, xb
    variable lin_pred5b already defined
    r(110);

    end of do-file

    r(110);

148 . do "C:\Users\cscho\AppData\Local\Temp\STD1a38_000000.tmp"

149 . pmcalplot prob5b los5, ci keep name(calibration_plot, replace) xtitle("Predicted probability", size(medsmall)) ytitle
    > size(medsmall)) legend(off)
    Binary option selected: Calibration plot for logistic prediction model displaying...
    Variable with name 'obs_pmcalplot' already exists, pmcalplot cannot generate required variables
    Variable with name 'exp_pmcalplot' already exists, pmcalplot cannot generate required variables
    Variable with name 'groups_pmcalplot' already exists, pmcalplot cannot generate required variables

150 .
151 . graph save "calibration_plot" "G:\My Drive\EBMA\Client Drive\QEII Jubilee\EBMA Working\Publications\Predicting extend
    > bration_plot - Model Updatedv2.gph", replace
    file G:\My Drive\EBMA\Client Drive\QEII Jubilee\EBMA Working\Publications\Predicting extended stay\Update2022\calibrati
    > v2.gph saved

152 .
153 . graph export "G:\My Drive\EBMA\Client Drive\QEII Jubilee\EBMA Working\Publications\Predicting extended stay\Update202
    > del Updatedv2.png", as(png) name("calibration_plot") replace
    file G:\My Drive\EBMA\Client Drive\QEII Jubilee\EBMA Working\Publications\Predicting extended stay\Update2022\calibrati
        Updatedv2.png saved as PNG format

154 .
155 . graph close

156 .
    end of do-file

157 . do "C:\Users\cscho\AppData\Local\Temp\STD1a38_000000.tmp"

158 . *Discrimination
159 . roctab los5 prob5b

                          ROC                    Asymptotic normal
               Obs       area     Std. err.     [95% conf. interval]
                     ------------------------------------------------
               200     0.6880       0.0394       0.61073      0.76535

160 .
161 . *Alternative
162 . brier los5 prob5b

    Mean probability of outcome     0.3150
                     of forecast    0.3150

    Correlation                     0.2961
    ROC area                        0.6880 p =   0.0000
```

| | |
|---|---|
| Brier score | **0.1969** |
| Spiegelhalter's z-statistic | **0.0489** p = **0.4805** |
| Sanders-modified Brier score | **0.1966** |
| Sanders resolution | **0.1867** |
| Outcome index variance | **0.2158** |
| Murphy resolution | **0.0291** |
| Reliability-in-the-small | **0.0099** |
| Forecast variance | **0.0196** |
| Excess forecast variance | **0.0178** |
| Minimum forecast variance | **0.0017** |
| Reliability-in-the-large | **0.0000** |
| 2*Forecast-Outcome-Covar | **0.0385** |

163 .
164 . *E/O
165 . summ prob5b

| Variable | Obs | Mean | Std. dev. | Min | Max |
|---|---|---|---|---|---|
| prob5b | 200 | .315 | .1402225 | .0895499 | .7599661 |

166 . summ los5

| Variable | Obs | Mean | Std. dev. | Min | Max |
|---|---|---|---|---|---|
| los5 | 230 | .3304348 | .4713956 | 0 | 1 |

167 .
168 . * check the expected and observed probabilities for the 10th risk group
169 . su obs_pmcalplot exp_pmcalplot if groups_pmcalplot==10

| Variable | Obs | Mean | Std. dev. | Min | Max |
|---|---|---|---|---|---|
| obs_pmcalp~t | **20** | **.6** | **0** | **.6** | **.6** |
| exp_pmcalp~t | **20** | **.9607756** | **0** | **.9607756** | **.9607756** |

170 .
171 . ***Conduct decision curve analysiss***
172 .
173 . dca los5 prob5b, probability(yes) smooth xstop(0.75) lcolor(black gs8 black) ///
> lpattern(solid solid dash) title("Decision Curve Analysis Updated Model", ///
>       size(4) color(red)) scheme(s1mono) saving(dcaresults,
replace) file **dcaresults.dta** saved

174 .
175 . graph save "Graph" "G:\My Drive\EBMA\Client Drive\QEII Jubilee\EBMA Working\Publications\Predicting extended stay\Upd
> Analysisv2.gph", replace
file **G:\My Drive\EBMA\Client Drive\QEII Jubilee\EBMA Working\Publications\Predicting extended stay\Update2022\Decision**
> ved

176 .
177 . graph export "G:\My Drive\EBMA\Client Drive\QEII Jubilee\EBMA Working\Publications\Predicting extended stay\Update202
> 2.png", as(png) name("Graph") replace
file **G:\My Drive\EBMA\Client Drive\QEII Jubilee\EBMA Working\Publications\Predicting extended stay\Update2022DecisionCu**
saved as PNG format

178 .
179 . graph close

180 .
end of do-file

181 .
